# Changes in cardiovascular disease monitoring in English primary care during the COVID-19 pandemic: an observational cohort study

**DOI:** 10.1101/2020.12.11.20247742

**Authors:** Clare R Bankhead, Sarah Lay-Flurrie, Brian D Nicholson, James P Sheppard, Chris P Gale, Harshana Liyanage, Dylan McGagh, Mark Minchin, Rafael Perera, Julian Sherlock, Margaret Smith, Nicholas PB Thomas, Cynthia Wright Drakesmith, Simon de Lusignan, FD Richard Hobbs

**Affiliations:** Nuffield Department of Primary Care Health Sciences, University of Oxford, Oxford, UK; NIHR Oxford Biomedical Research Centre, Oxford University Hospitals NHS Foundation Trust; Leeds Institute of Cardiovascular and Metabolic Medicine, University of Leeds, Leeds, UK; Leeds Institute for Data Analytics, University of Leeds, Leeds, UK; Department of Cardiology, Leeds Teaching Hospitals NHS Trust, Leeds, UK; Magdalen College, University of Oxford, OX1 4AU; National Institute for Health and Care Excellence (NICE). Level 1A, City Tower, Piccadilly Plaza, Manchester, M1 4BT; Royal College of General Practitioners, 30 Euston Square, London, NW1 2FB

## Abstract

**Objective:** To quantify the impact and recovery in cardiovascular disease monitoring in primary care associated with the first COVID-19 lockdown.

**Design:** Retrospective nationwide primary care cohort study, utilising data from 1st January 2018 to 27^th^ September 2020.

**Setting:** We extracted primary care electronic health records data from 514 primary care practices in England contributing to the Oxford Royal College of General Practitioners Clinical Informatics Digital Hub (ORCHID). These practices were representative of English primary care across urban and non-urban practices.

**Participants:** The ORCHID database included 6,157,327 active patients during the study period, and 13,938,390 patient years of observation (final date of follow-up 27^th^ September 2020). The mean (SD) age was 38±24 years, 49.4% were male and the majority were of white ethnicity (65% [21.9% had unknown ethnicity])

**Exposure:** The primary exposure was the first national lockdown in the UK, starting on 23^rd^ March 2020.

**Main outcome measures:** Records of cholesterol, blood pressure, HbA1c and International Normalised Ratio (INR) measurement derived from coded entries in the primary care electronic health record.

**Results:** Rates of cholesterol, blood pressure, HbA1c and INR recording dropped by 23-87% in the week following the first UK national lockdown, compared with the previous week. The largest decline was seen in cholesterol (IRR 0.13, 95% CI 0.11 to 0.15) and smallest for INR (IRR 0.77, 95% CI 0.72 to 0.81).

Following the immediate drop, rates of recorded tests increased on average by 5-9% per week until 27^th^ September 2020. However, the number of recorded measures remained below that expected for the time of year, reaching 51.8% (95% CI 51.8 to 51.9%) for blood pressure, 63.7%, (95% CI 63.7% to 63.8%) for cholesterol measurement and 70.3% (95% CI 70.2% to 70.4%) for HbA1c. Rates of INR recording declined throughout the previous two years, a trend that continued after lockdown. There were no differences in the times series trends based on sex, age, ethnicity or deprivation.

**Conclusions:** Cardiovascular disease monitoring in English primary care declined substantially from the time of the first UK lockdown. Despite a consistent recovery in activity, there is still a substantial shortfall in the numbers of recorded measurements to those expected. Strategies are required to ensure cardiovascular disease monitoring is maintained during the COVID-19 pandemic.

## Introduction

The Coronavirus Disease 2019 (COVID-19) pandemic, and associated restrictions on daily life are thought to have dramatically affected access to healthcare across the world. In the UK, from early March 2020, any patients with COVID-like symptoms were advised to call NHS 111 for advice, rather than to seek care in-person. In addition, telephone and video triage was recommended across primary care in order to reduce the number of face-to-face consultations.^1^ A million and a half people in England who were considered extremely vulnerable to COVID-19 were advised to ‘shield’ at home for at least 12 weeks.^2^ Following the COVID-19 lockdown on 23^rd^ March 2020, there was an almost ubiquitous shift to remote consultations, as healthcare settings developed “COVID secure” ways of working.^3-5^ Many routine appointments, procedures and non-urgent care, in both primary and secondary care, were cancelled to divert resources to emergency care for people with suspected or confirmed COVID-19.^6,7^ Early data showed that endoscopy procedures reduced by 95%,^8^ cancer referrals dropped by 70%,^9^ hospital admissions with stroke and acute myocardial infarction decreased by 50%,^10,11^ attendances at accident and emergency dropped by 35%^12^ and routine safety monitoring tests for psychiatric medications became difficult to obtain.^13^ In primary care, data are limited but one study of 47 practices in a single health region in England reported a halving in the number of diagnoses of type 2 diabetes, and anxiety and depression, in primary care and a 43% drop in the diagnosis of circulatory conditions.^14^ It is uncertain whether this was reflected across the country, or specific to northern England where COVID-19 infection rates are thought to have been higher.^15^

Early data from hospital admissions for acute coronary syndrome in England showed that following an initial 40% reduction at the start of the UK lockdown, a recovery in admission rates was observed to the end of May 2020, but remained 16% lower than expected.^6^ It is currently unclear as to whether routine primary and secondary care activity has recovered to pre-pandemic levels as lockdown restrictions were eased. A sustained drop in activity in primary care is clinically important. The majority of people with a new diagnosis of serious disease first attend primary care^16^ and are then referred. Most major disease prevention is conducted in primary care, for example risk factor assessment to focus management on those at highest risk, and the monitoring of chronic disease to reduce subsequent morbidity and excess mortality. In particular, cardiovascular disease prevention programmes are reliant on the routine measurement of risk factors such as cholesterol, blood pressure and glycated haemoglobin (HbA1c) for both diagnosis, treatment initiation and risk factor control.

Understanding how primary care activity was affected by the first UK lockdown may help avert this reduced primary care activity in the first wave of COVID-19 translating into excess mortality in the future. Crucially, data are required that describe whether primary care provision for non-COVID conditions such as cardiovascular disease recovered following the first lockdown. This mayenable the development of strategies to mitigate persistent shortfalls in key clinical areas. Furthermore, quantifying this key role of general practice, namely the delivery of disease prevention, during the COVID-19 pandemic is also important given the highly critical concerns expressed by some commentators that general practice has failed the public during COVID. ^17^ The present study analysed the rates of routine cardiovascular disease monitoring in primary care across the past three years to quantify the impact and recovery in clinical care associated with the first COVID-19 lockdown.

## Methods

### Design

We conducted a retrospective nationwide cohort study, utilising electronic health records from primary care practices in England contributing to the Oxford Royal College of General Practitioners Clinical Informatics Digital Hub (ORCHID).^18^ The participating practices were representative of English primary care across urban and non-urban practices.^19^ Two experienced PPI participants commented on the protocol and the research aims. The protocol for this study was accepted by an independent approval committee and received ethical approval from the University of Oxford, Medical Sciences Interdivisional Research Ethics Committee (ref: R69874/RE001).

### Study population, exposure, and outcomes

This study included all registered patients of any age within ORCHID. Data were extracted for all consultations occurring between 1^st^ January 2018 and 27^th^ September 2020. Person years of observation were calculated and used as the denominator for all analyses.

The primary exposure of interest was the date of national lockdown in the UK, 23^rd^ March 2020 (figure 1).

**Figure 1.**
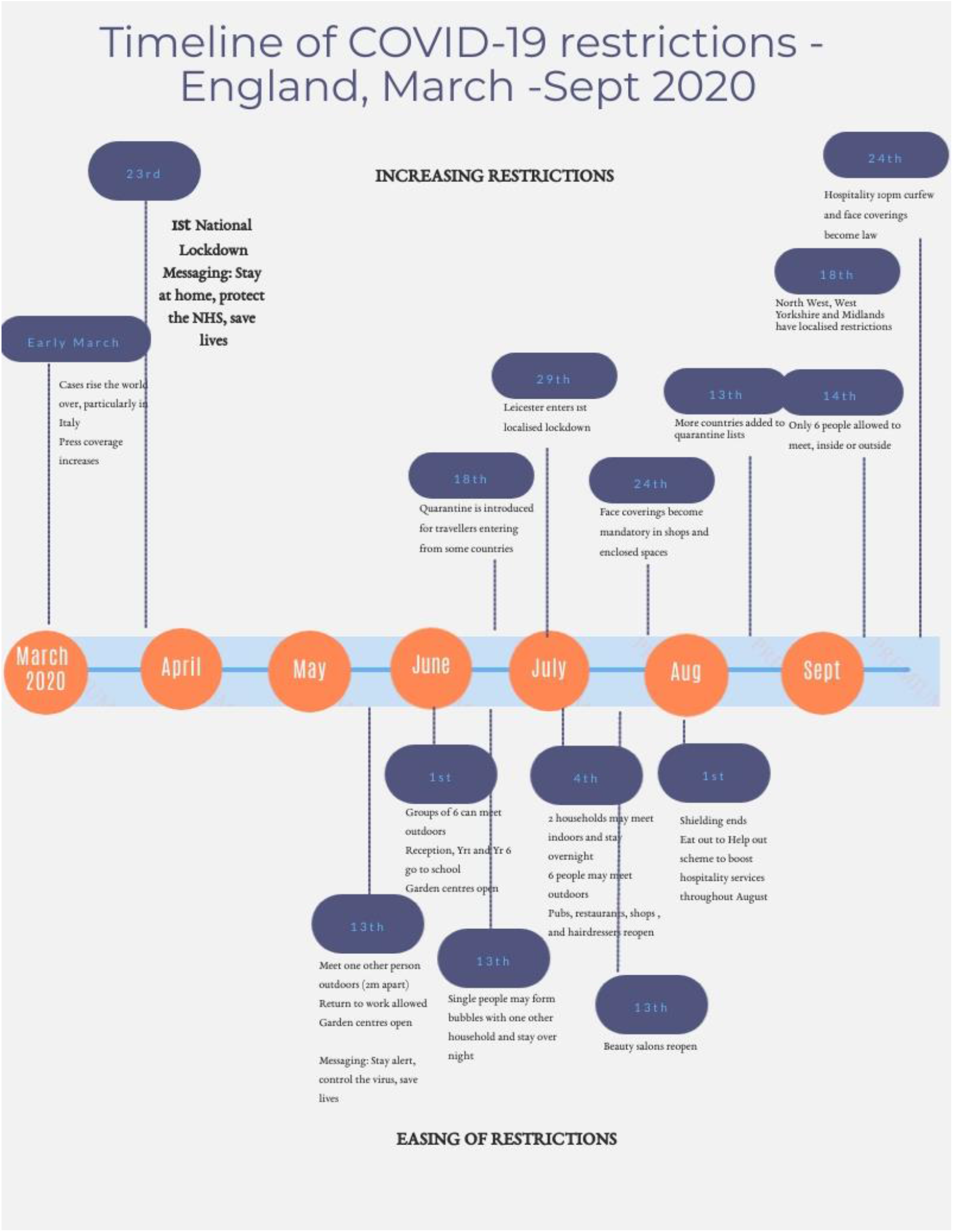
Timeline of UK lockdowns and restrictions

The cardiovascular disease monitoring outcomes included records of cholesterol, blood pressure (BP), HbA1c, and International Normalised Ratio (INR) measurement. All outcomes were based on codes entered into the primary care electronic health record and were curated in SNOMED CT.(complete variable lists are included in appendix 1).

### Covariates

Covariates were age group (<15, 15-24, 25-44, 45-64, 65-74, 75-84 and 85+), sex, ethnicity, and Indices of Multiple Deprivation (IMD quintile). Those with missing ethnicity were classed as unknown, and the small number with missing IMD data were excluded (174,004; 2.8%) from stratified analyses involving IMD.

### Statistical analysis

Descriptive statistics were used to define the characteristics of the study population. Weekly rates of cardiovascular disease monitoring were calculated and plotted over time from 1^st^ January 2018 through to 27^th^ September 2020, overall and stratified by covariate groups.

Negative binomial regression models, including a term for the week of follow-up and an indicator term for before/after lockdown were fitted to examine the rates of testing during the study period. Incident rate ratios (IRR) and 95% confidence intervals (CI) were derived from these models to compare clinical activity rates in the week after lockdown, compared with the week prior to lockdown.

Estimates of the rate of change in clinical activity each week after the start of lockdown were also obtained from the models. For each outcome, five models were fitted in total: one model for the overall rates as described and one model for rates stratified by each of the four covariates (age, sex, ethnicity and IMD).

To estimate whether the effect of lockdown and subsequent recovery varied by covariate groups, the latter models included additional terms for the covariate, interactions between the covariate and week of follow-up and the covariate and lockdown indicator (equivalent to a stratified analysis).

Expected number of tests in each week were predicted based on observed rates in 2018 and 19, standardised to the 2020 denominator. Expected and actual number of tests in 2020 were summed from January to 27^th^ September and the cumulative sum displayed graphically. The recovery in cumulative activity was expressed as the proportion of actual tests recorded in 2020 compared to the average expected number over the same time period in 2018 and 2019. Analyses were conducted using R (version 3.5.3).

## Results

The ORCHID database included 6,157,327 active patients during the study period, from 514 general practices. Included patients had a mean age of 38±sd 24 years, 49.4% were male and the majority were of white ethnicity (65% [21.9% had unknown ethnicity]) (table 1). In total, there were 13,938,390 patient years of observation, and the vast majority of patients were included for the entire study period (median period of observation of 2.74 years, IQR 1.94 to 2.74 years).

**Table 1.** Characteristics of patients included in the analysis

| Patient characteristic | Total population<br>(n=6,157,327) |  |
| --- | --- | --- |
|  | Mean/N | SD/% |
| **Male** | 3,040,122 | 49.4% |
| **Age (years)** | 38.4 | 23.7 |
| **Ethnic group** |  |  |
| **White** | 3,994,676 | 64.9% |
| **Asian** | 441,621 | 7.2% |
| **Black** | 190,518 | 3.1% |
| **Mixed** | 100,595 | 1.6% |
| **Other** | 82,325 | 1.3% |
| **Unknown** | 1,347,592 | 21.9% |
| **IMD Quintile** |  |  |
| **1 (most deprived)** | 1,064,168 | 17.3% |
| **2** | 1,136,632 | 18.5% |
| **3** | 1,175,897 | 19.1% |
| **4** | 1,214,756 | 19.7% |
| **5** | 1,391,870 | 22.6% |
| **Unknown** | 174,004 | 2.8% |
| **Urban/ rural** |  |  |
| **Urban** | 1,032,343 | 16.8% |
| **Rural** | 5,097,545 | 82.8% |

IMD= Indices of Multiple Deprivation

Figure 2 shows the rate of cholesterol, blood pressure, HbA1c and INR recording across the study period. The incidence dropped significantly by 23-87% in the week that the UK went into national lockdown, compared to the week before (table 2). The change in the incidence rate (in the week before and after lockdown) was largest for cholesterol (IRR 0.13, 95% CI 0.11 to 0.15) and smallest for INR (IRR 0.77, 95% CI 0.72 to 0.81). There were no differences in the rates based on sex, age, ethnicity or socioeconomic deprivation (table 2).

**Table 2.** Incidence rate of test activity the week after/before UK lockdown by patient characteristics

| Characteristic | Cholesterol |  |  | Blood pressure |  |  | HbA1c |  |  | INR |  |  |
| --- | --- | --- | --- | --- | --- | --- | --- | --- | --- | --- | --- | --- |
|  | IRR | 95% CI |  | IRR | 95% CI |  | IRR | 95% CI |  | IRR | 95% CI |  |
| **Overall** | 0.13 | 0.11 | - 0.15 | 0.17 | 0.15 | - 0.19 | 0.16 | 0.14 | - 0.19 | 0.77 | 0.72 | - 0.81 |
| **Sex** |  |  |  |  |  |  |  |  |  |  |  |  |
| Female | 0.12 | 0.10 | - 0.14 | 0.17 | 0.15 | - 0.19 | 0.16 | 0.13 | - 0.18 | 0.75 | 0.70 | - 0.79 |
| Male | 0.14 | 0.12 | - 0.17 | 0.17 | 0.15 | - 0.20 | 0.17 | 0.14 | - 0.20 | 0.78 | 0.74 | - 0.83 |
| **Age group (years)** |  |  |  |  |  |  |  |  |  |  |  |  |
| <15 | 0.29 | 0.23 | - 0.36 | 0.11 | 0.10 | - 0.13 | 0.16 | 0.13 | - 0.19 | 0.48 | 0.40 | - 0.57 |
| 15-24 | 0.20 | 0.16 | - 0.24 | 0.15 | 0.13 | - 0.17 | 0.20 | 0.17 | - 0.24 | 0.64 | 0.56 | - 0.72 |
| 25-44 | 0.13 | 0.11 | - 0.16 | 0.17 | 0.15 | - 0.19 | 0.17 | 0.15 | - 0.20 | 0.71 | 0.66 | - 0.76 |
| 45-64 | 0.13 | 0.11 | - 0.16 | 0.15 | 0.13 | - 0.17 | 0.16 | 0.14 | - 0.19 | 0.81 | 0.76 | - 0.86 |
| 65-74 | 0.12 | 0.10 | - 0.14 | 0.17 | 0.15 | - 0.19 | 0.15 | 0.12 | - 0.17 | 0.78 | 0.73 | - 0.83 |
| 75-84 | 0.12 | 0.10 | - 0.14 | 0.19 | 0.17 | - 0.22 | 0.16 | 0.13 | - 0.19 | 0.75 | 0.71 | - 0.80 |
| 85+ | 0.12 | 0.10 | - 0.14 | 0.27 | 0.23 | - 0.30 | 0.18 | 0.15 | - 0.22 | 0.77 | 0.73 | - 0.83 |
| **Ethnic group** |  |  |  |  |  |  |  |  |  |  |  |  |
| White | 0.13 | 0.11 | - 0.15 | 0.17 | 0.15 | - 0.19 | 0.16 | 0.14 | - 0.19 | 0.77 | 0.73 | - 0.82 |
| Black | 0.13 | 0.12 | - 0.16 | 0.17 | 0.15 | - 0.20 | 0.16 | 0.13 | - 0.19 | 0.82 | 0.73 | - 0.92 |
| Asian | 0.12 | 0.11 | - 0.14 | 0.15 | 0.13 | - 0.17 | 0.14 | 0.12 | - 0.16 | 0.73 | 0.66 | - 0.80 |
| Mixed | 0.16 | 0.14 | - 0.19 | 0.17 | 0.15 | - 0.20 | 0.18 | 0.15 | - 0.22 | 0.73 | 0.61 | - 0.89 |
| Other | 0.13 | 0.11 | - 0.15 | 0.16 | 0.14 | - 0.18 | 0.15 | 0.12 | - 0.18 | 0.95 | 0.80 | - 1.12 |
| Unknown | 0.15 | 0.13 | - 0.17 | 0.17 | 0.15 | - 0.20 | 0.18 | 0.15 | - 0.21 | 0.76 | 0.71 | - 0.81 |
| **IMD Quintile** |  |  |  |  |  |  |  |  |  |  |  |  |
| 1 (most deprived) | 0.12 | 0.10 | - 0.14 | 0.16 | 0.14 | - 0.18 | 0.15 | 0.13 | - 0.18 | 0.76 | 0.72 | - 0.81 |
| 2 | 0.13 | 0.11 | - 0.15 | 0.17 | 0.15 | - 0.19 | 0.16 | 0.14 | - 0.19 | 0.76 | 0.71 | - 0.81 |
| 3 | 0.13 | 0.11 | - 0.16 | 0.17 | 0.15 | - 0.19 | 0.17 | 0.14 | - 0.20 | 0.78 | 0.73 | - 0.83 |
| 4 | 0.13 | 0.11 | - 0.16 | 0.17 | 0.15 | - 0.20 | 0.16 | 0.14 | - 0.19 | 0.78 | 0.73 | - 0.83 |
| 5 | 0.13 | 0.11 | - 0.15 | 0.17 | 0.15 | - 0.20 | 0.16 | 0.14 | - 0.19 | 0.76 | 0.72 | - 0.81 |

IRR=incidence rate ratio (the week after UK lockdown on 23<sup>rd</sup> March 2020 vs the week before)

**Figure 2.**
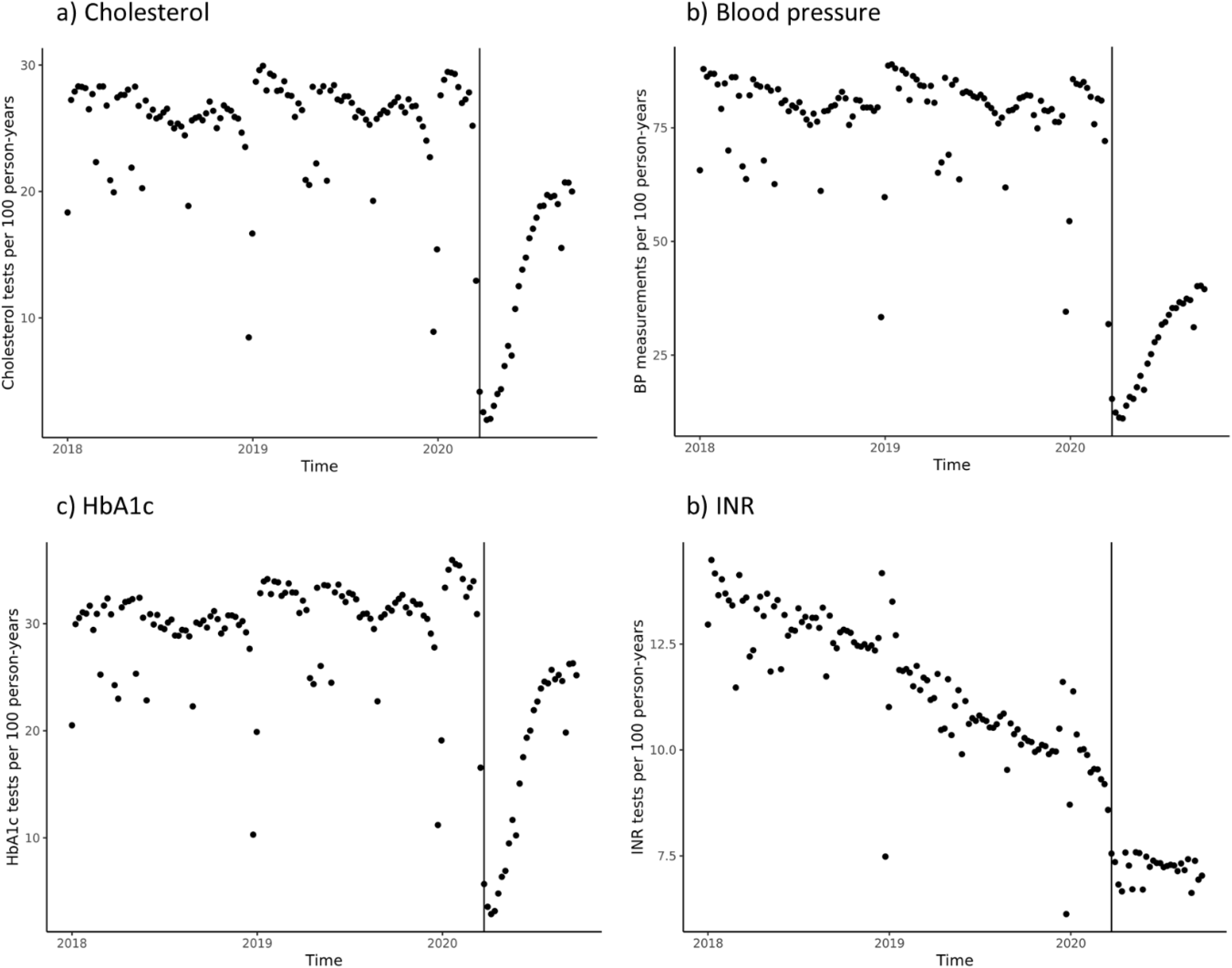
Rates of testing (per week) for cholesterol, blood pressure, HbA1c and INR across primary care from 1^st^ January 2018 to 27^st^ September 2020

Following the immediate decline after lockdown began, rates of monitoring for cholesterol, blood pressure and HbA1c increased, on average, by 5-9% per week until 27^th^ September (table 3), but did not recover to pre-lockdown levels. Compared with the same week in 2019, recording rates during the last week of September 2020 were lowest for blood pressure at 48.4% (95% CI 47.9% to 49.1%), 76.2% (95% 74.8% to 77.6%) for cholesterol, and 80.7% (95% CI 79.3% to 82.0%) for HbA1c of the previous year’s rates. There was no such increase for INR following the initial drop observed immediately after lockdown (IRR 0.999, 95% CI 0.996 to 1.003; table 3). There were no differences in rates following lockdown across patient characteristics (table 3).

**Table 3.**
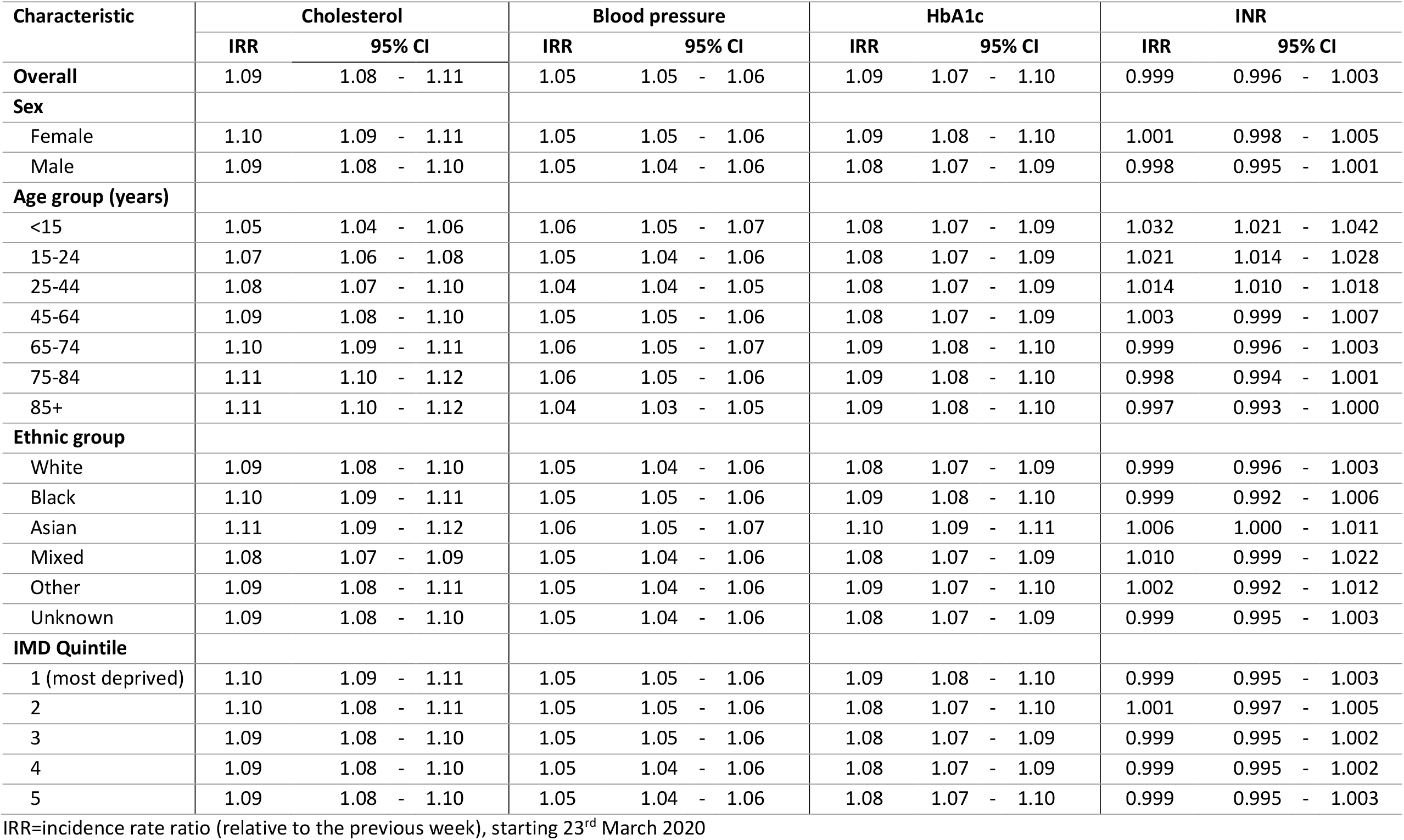
Incidence rate per week of test activity during recovery after UK lockdown by patient characteristics

| Characteristic | Cholesterol |  |  | Blood pressure |  |  | HbA1c |  |  | INR |  |  |
| --- | --- | --- | --- | --- | --- | --- | --- | --- | --- | --- | --- | --- |
|  | IRR | 95% CI |  | IRR | 95% CI |  | IRR | 95% CI |  | IRR | 95% CI |  |
| **Overall** | 1.09 | 1.08 | - 1.11 | 1.05 | 1.05 | - 1.06 | 1.09 | 1.07 | - 1.10 | 0.999 | 0.996 | - 1.003 |
| **Sex** |  |  |  |  |  |  |  |  |  |  |  |  |
| Female | 1.10 | 1.09 | - 1.11 | 1.05 | 1.05 | - 1.06 | 1.09 | 1.08 | - 1.10 | 1.001 | 0.998 | - 1.005 |
| Male | 1.09 | 1.08 | - 1.10 | 1.05 | 1.04 | - 1.06 | 1.08 | 1.07 | - 1.09 | 0.998 | 0.995 | - 1.001 |
| **Age group (years)** |  |  |  |  |  |  |  |  |  |  |  |  |
| <15 | 1.05 | 1.04 | - 1.06 | 1.06 | 1.05 | - 1.07 | 1.08 | 1.07 | - 1.09 | 1.032 | 1.021 | - 1.042 |
| 15-24 | 1.07 | 1.06 | - 1.08 | 1.05 | 1.04 | - 1.06 | 1.08 | 1.07 | - 1.09 | 1.021 | 1.014 | - 1.028 |
| 25-44 | 1.08 | 1.07 | - 1.10 | 1.04 | 1.04 | - 1.05 | 1.08 | 1.07 | - 1.09 | 1.014 | 1.010 | - 1.018 |
| 45-64 | 1.09 | 1.08 | - 1.10 | 1.05 | 1.05 | - 1.06 | 1.08 | 1.07 | - 1.09 | 1.003 | 0.999 | - 1.007 |
| 65-74 | 1.10 | 1.09 | - 1.11 | 1.06 | 1.05 | - 1.07 | 1.09 | 1.08 | - 1.10 | 0.999 | 0.996 | - 1.003 |
| 75-84 | 1.11 | 1.10 | - 1.12 | 1.06 | 1.05 | - 1.06 | 1.09 | 1.08 | - 1.10 | 0.998 | 0.994 | - 1.001 |
| 85+ | 1.11 | 1.10 | - 1.12 | 1.04 | 1.03 | - 1.05 | 1.09 | 1.08 | - 1.10 | 0.997 | 0.993 | - 1.000 |
| **Ethnic group** |  |  |  |  |  |  |  |  |  |  |  |  |
| White | 1.09 | 1.08 | - 1.10 | 1.05 | 1.04 | - 1.06 | 1.08 | 1.07 | - 1.09 | 0.999 | 0.996 | - 1.003 |
| Black | 1.10 | 1.09 | - 1.11 | 1.05 | 1.05 | - 1.06 | 1.09 | 1.08 | - 1.10 | 0.999 | 0.992 | - 1.006 |
| Asian | 1.11 | 1.09 | - 1.12 | 1.06 | 1.05 | - 1.07 | 1.10 | 1.09 | - 1.11 | 1.006 | 1.000 | - 1.011 |
| Mixed | 1.08 | 1.07 | - 1.09 | 1.05 | 1.04 | - 1.06 | 1.08 | 1.07 | - 1.09 | 1.010 | 0.999 | - 1.022 |
| Other | 1.09 | 1.08 | - 1.11 | 1.05 | 1.04 | - 1.06 | 1.09 | 1.07 | - 1.10 | 1.002 | 0.992 | - 1.012 |
| Unknown | 1.09 | 1.08 | - 1.10 | 1.05 | 1.04 | - 1.06 | 1.08 | 1.07 | - 1.09 | 0.999 | 0.995 | - 1.003 |
| **IMD Quintile** |  |  |  |  |  |  |  |  |  |  |  |  |
| 1 (most deprived) | 1.10 | 1.09 | - 1.11 | 1.05 | 1.05 | - 1.06 | 1.09 | 1.08 | - 1.10 | 0.999 | 0.995 | - 1.003 |
| 2 | 1.10 | 1.08 | - 1.11 | 1.05 | 1.05 | - 1.06 | 1.08 | 1.07 | - 1.10 | 1.001 | 0.997 | - 1.005 |
| 3 | 1.09 | 1.08 | - 1.10 | 1.05 | 1.05 | - 1.06 | 1.08 | 1.07 | - 1.09 | 0.999 | 0.995 | - 1.002 |
| 4 | 1.09 | 1.08 | - 1.10 | 1.05 | 1.04 | - 1.06 | 1.08 | 1.07 | - 1.09 | 0.999 | 0.995 | - 1.002 |
| 5 | 1.09 | 1.08 | - 1.10 | 1.05 | 1.04 | - 1.06 | 1.08 | 1.07 | - 1.10 | 0.999 | 0.995 | - 1.003 |

IRR=incidence rate ratio (relative to the previous week), starting 23<sup>rd</sup> March 2020

Overall, recording of cardiovascular disease risk factors was significantly reduced compared with previous years (figure 3). By the 27^th^ September 2020, there were 355,139 fewer records for cholesterol, (63.7% of expected, 95% CI 63.7% to 63.8%); 1,442,792 fewer BP records (51.8% of expected, 95% CI 51.8% to 51.9%); 337,659 fewer HbA1c records (70.3% of expected, 95% CI 70.2%to 70.4%); and 355,138 fewer INR records (63.7% of expected, 95% CI 63.7% to 63.8%).

**Figure 3.**
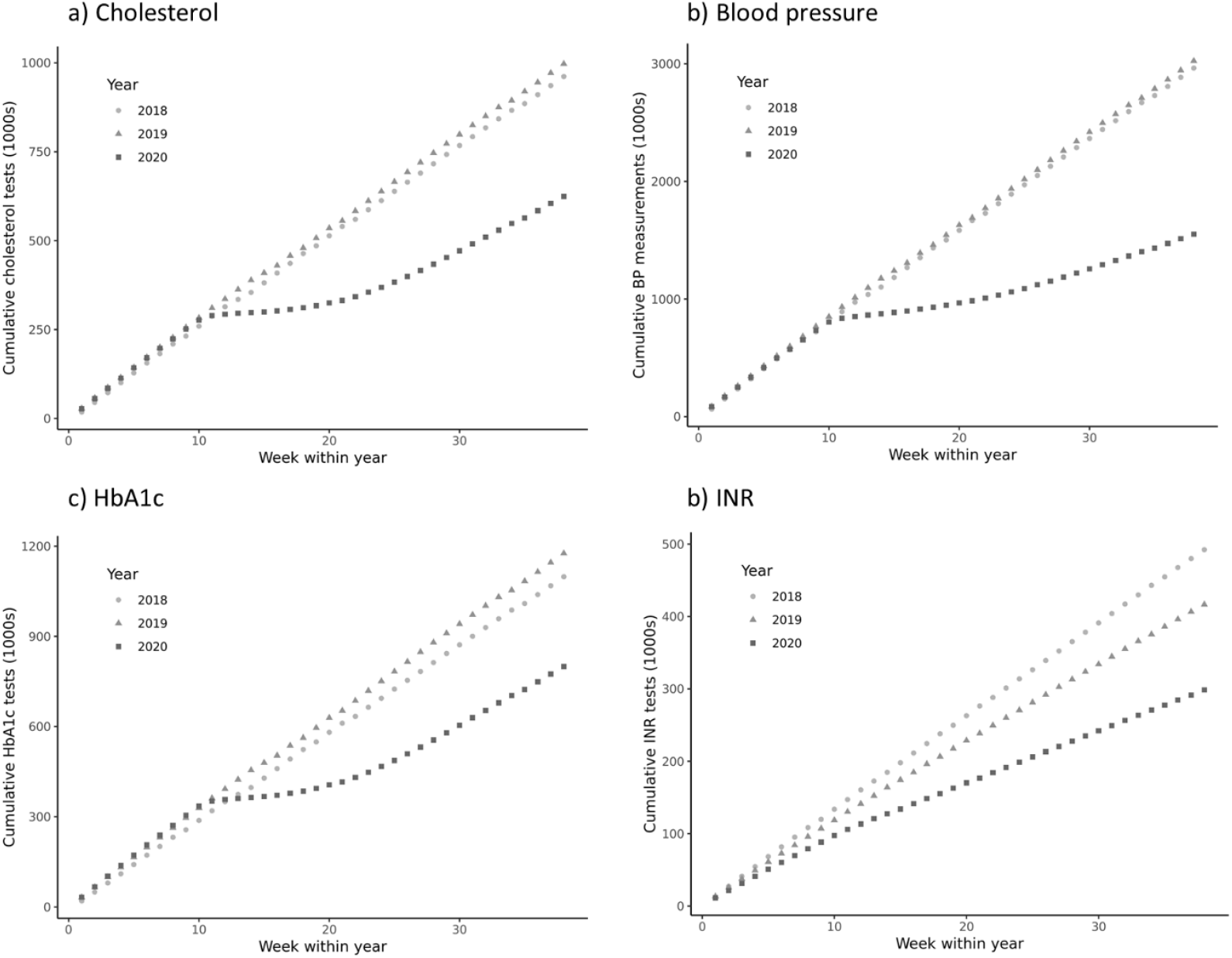
Cumulative rates of testing (per week) for cholesterol, blood pressure, HbA1c and INR across primary care by year (from 2018 to 2020)

## Discussion

### Summary of main findings

In a nationally representative cohort of 6,157,327 registered patients, cholesterol, blood pressure, HbA1c and INR recording, all measures that require in person consulting, were found to be reduced by up to 87% following the UK government’s decision to enter into a national lockdown at the end of March 2020. Rates of cholesterol, blood pressure and HbA1c recording subsequently increased by 5-9% per week, although not to pre-lockdown levels, reaching between 52% and 70% of the expected number based on the previous years. Overall, there were between 355,138 and 1,442,792 fewer tests (depending on the measure) conducted by the end of September 2020 compared with the previous years. No such recovery was observed for INR testing, possibly due to many patients being switched from warfarin (which requires INR monitoring) to direct oral anticoagulants (which do not), which had been occurring prior to the pandemic^20^, but which was accelerated to avoid unnecessary patient contact with primary care during the pandemic.^21,22^

### Strengths and limitations

This is the first analysis of nationally representative data from primary care in England showing both the immediate drop in cardiovascular disease monitoring and subsequent recovery during the COVID-19 pandemic. Data were derived from the ORCHID database which is capable of weekly data downloads, permitting some of the most timely and up-to-date analyses of primary care data in the world. Analyses are limited to those coded in patients’ electronic health records. It is possible that changes in clinical practice, such as the switch to remote consultations ^1,3,4^ or reduced administrative support for general practitioners may have affected coding of risk factor measurement. In addition, some patients may have continued to monitor their cardiovascular risk factors remotely through self-monitoring, ^23-25^ but this may not have been captured in the electronic health record.

We focussed on cardiovascular disease since this is responsible for most premature deaths and major morbidity In the UK, but there are major disease prevention strategies possible with the focus on the early detection of risk factors and the close management of these risk factors monitored by follow up testing. General practice is largely responsible for this major public health delivery.

This was an observational study and analyses were not designed to infer causality between the initiation of a national lockdown and subsequent cardiovascular disease monitoring. In addition, due to the contemporary nature of the data, it was not possible to examine the association between changes in clinical activity and subsequent clinical outcomes such as cardiovascular morbidity and mortality. This will become possible in time but, at present, these analyses highlight the scale of reduced cardiovascular disease monitoring in UK primary care.

### Comparison with existing literature

To date, the majority of existing studies examining the impact of the COVID-19 pandemic on non-COVID-19 related clinical care have focussed on secondary care activity. These studies have shown decreases in hospital attendances and admissions ranging between 35% for accident and emergency attendance^12^ and 95% for endoscopy procedures^8^. Admissions for acute myocardial infarctions dropped by 42%^11^ and cardiac procedures were reduced by around 50%^7^. There is some evidence of recovery in secondary care with admissions for acute coronary syndrome, including myocardial infarction, returning to 16% below the previous year average by the end of May.^6^ Similar data have been reported in cardiovascular services in Denmark ^26^ and the United States of America^27^. Secondary care activity has been reported to have decrease globally for urology,^28^ paediatric rheumatology,^29^ and gynaecological oncology.^30^

In primary care, one small study of 47 urban general practices contributing data to the Salford Integrated Record database found the number of diagnoses of anxiety and depression, type 2 diabetes and circulatory conditions fell by 43%-50% in the period between March and May 2020.^14^

The present analysis examined data from 6,157,327 patients from 514 general practices across England and focussed on cardiovascular disease. We found larger reductions in clinical activity of up to 83% for cholesterol, blood pressure and HbA1c recording, most likely due to these tests requiring face-to-face appointment in order to record them.

We also observed an almost immediate subsequent increase in cardiovascular disease related activity which continued to increase by 5-9% per week throughout the rest of the study period. This is the first study to quantify the recovery of primary care activity since the start of UK lockdown in any clinical area. Although this recovery was rapid, rates of clinical activity did not return to pre-lockdown levels leaving a substantial and persistent deficit of ‘missed’ records, compared to previous years.

Whilst the reduction in testing and failure to fully recover to pre-lockdown levels may reflect improved efficiency of service provision, with only the sickest patients being tested, such a large drop in overall activity could have a material impact on cardiovascular morbidity and excess mortality and should therefore be mitigated when possible.

The reduction in recorded cardiovascular disease monitoring is almost certainly the result of a combination of altered patient behaviour, changes in access to primary care, modifications in clinical practice, as well as an excess in deaths.^31^ Fear of contracting COVID-19 from healthcare settings and shielding advice for older and comorbid people to remain at home has influenced help-seeking. Understanding how to balance and communicate the risks of reduced cardiovascular disease monitoring with the risks of COVID-19 exposure in community healthcare settings will be vital to adequately reinstate care to those most vulnerable.

Remote consultation may have resulted in fewer opportunistic blood pressure measurements when patients attend for a non-blood pressure related problem. Further analyses are required to determine whether reductions in activity are associated with reductions in tests used for diagnosis compared to those used for monitoring.

Recorded testing for cholesterol and HbA1c were conducted much less frequently pre-lockdown than blood pressure (∼30 tests per 100 patient years vs. ∼85 blood pressure measurements per 100 person years), therefore a smaller reduction in cumulative post-lockdown testing would be expected in less frequent measurements, given the relatively short duration since lockdown. By contrast, INR testing was already in decline from 2018-2019 and this was amplified at the start of lockdown, presumably due to an acceleration of switching patients from warfarin to direct oral anticoagulants that do not require regular blood test monitoring.^20,21^

Reduced recording may represent decreased monitoring and it has been shown that blood pressure control is obtained more quickly with frequent monitoring,^32,33^ so a lack of measurement may increase adverse events. However, many patients may be using home blood pressure monitors to safely manage their blood pressure and therefore safeguarding against harm. Based on previous modelling of vascular health checks programmes, routine measurement of cholesterol, blood pressure and HbA1c has the potential to reduce up to 1,300 major cardiovascular events per 100,000 people screened.^34^ Whether these events occur in the absence of cardiovascular disease monitoring depends on actions taken by general practitioners and policy makers now.

In response to the altered pressures in primary care, NHS England and NHS Improvement in agreement with the BMA’s General Practitioners Committee have simplified and reduced the Quality and Outcomes Framework requirements for 2020/21.^35^ It is possible that this could have a negative effect on the quality of recording in the electronic health records, and possibly on clinical care. ^36^

Given the importance of monitoring cardiovascular disease, and the substantial number of missing records this year, it is possible that the reduction in primary care activity for at least three of the four of the outcomes studied may result in increased cardiovascular disease morbidity and mortality if the usual control of these parameters has deteriorated. There is therefore an important need to develop approaches to identify whether cholesterol, blood pressure, and HbA1c control has been compromised as the pandemic continues. Early strategies, incorporating what we know about remote self-monitoring of risk factors,^23-25^ are required to address the shortfall in monitoring identified by this research. Although there were no apparent differences between testing rates for specific patient groups, further analyses are needed to determine whether certain tests, situations (e.g. diagnosis vs monitoring) or patient groups should be prioritised for these strategies.

### Conclusions

This analysis of a large nationally representative dataset from UK primary care shows both the immediate drop in cardiovascular disease monitoring and the subsequent recovery. Whilst there has been partial recovery of cardiovascular preventive activity in English general practice since lockdown, with consistent growth evident since the initial lockdown, reaching 51-68% of expected cumulative records. Importantly, there were no differences observed for the sudden reduction or subsequent recovery of preventive activity by patient characteristics, such as sex or ethnicity. However, there remains a substantial number of missing records. Given the importance of cardiovascular disease management in the prevention of premature death, it is possible that this shortfall in clinical activity will translate into increased cardiovascular disease morbidity and mortality. There is now an imperative to help primary care practices develop strategies to mitigate the decline in cardiovascular disease monitoring during and beyond the COVID-19 pandemic.

## Data Availability

Data were obtained from ORCHID

https://clininf.eu/index.php/orchid-data/

## Acknowledgements

Patients and practices in the Oxford-RCGP RSC network who allow data sharing.

## Conflicts of interest

Prof. Gale reports personal fees from Amgen, personal fees from AstraZeneca, personal fees from Bayer, personal fees from Daiichi Sankyo, personal fees from Vifor Pharma, grants from Abbot, grants from BMS, outside the submitted work.

All other authors have None to declare

