## Appendix 1 for "Changes in cardiovascular disease monitoring in English primary care during the COVID-19 pandemic: an observational cohort study"

| ConditionID | ConditionName | ConceptID | Term |
|---|---|---|---|
| 2051 | Blood Pressure - Both | 502541000000105 | [D]Blood pressure raised, hypertension not diagnosed (situation) |
| 2051 | Blood Pressure - Both | 502551000000108 | [D]Nonspecific low blood pressure reading (situation) |
| 2051 | Blood Pressure - Both | 207519004 | [D]Raised blood pressure reading (situation) |
| 2051 | Blood Pressure - Both | 314463006 | 24 hour blood pressure (observable entity) |
| 2051 | Blood Pressure - Both | 386534000 | Arterial blood pressure (observable entity) |
| 2051 | Blood Pressure - Both | 723232008 | Average blood pressure (observable entity) |
| 2051 | Blood Pressure - Both | 928031000000105 | Baseline blood pressure |
| 2051 | Blood Pressure - Both | 928021000000108 | Baseline blood pressure (observable entity) |
| 2051 | Blood Pressure - Both | 250765005 | Blood pressure (observable entity) |
| 2051 | Blood Pressure - Both | 75367002 | Blood pressure (observable entity) |
| 2051 | Blood Pressure - Both | 392570002 | Blood pressure finding (finding) |
| 2051 | Blood Pressure - Both | 314956000 | Borderline blood pressure (finding) |
| 2051 | Blood Pressure - Both | 170604000 | Borderline blood pressure (finding) |
| 2051 | Blood Pressure - Both | 147850007 | Borderline blood pressure (finding) |
| 2051 | Blood Pressure - Both | 1036541000000104 | Central blood pressure |
| 2051 | Blood Pressure - Both | 12763006 | Finding of decreased blood pressure (finding) |
| 2051 | Blood Pressure - Both | 24184005 | Finding of increased blood pressure (finding) |
| 2051 | Blood Pressure - Both | 62275004 | Hypertensive episode (disorder) |
| 2051 | Blood Pressure - Both | 251077004 | Invasive arterial pressure (observable entity) |
| 2051 | Blood Pressure - Both | 386532001 | Invasive arterial pressure (observable entity) |
| 2051 | Blood Pressure - Both | 386533006 | Invasive blood pressure (observable entity) |
| 2051 | Blood Pressure - Both | 163033001 | Lying blood pressure (observable entity) |
| 2051 | Blood Pressure - Both | 140246009 | Lying blood pressure reading (observable entity) |
| 2051 | Blood Pressure - Both | 852291000000105 | Maximum mean blood pressure (observable entity) |
| 2051 | Blood Pressure - Both | 6797001 | Mean blood pressure (observable entity) |
| 2051 | Blood Pressure - Both | 251076008 | Non-invasive arterial pressure (observable entity) |
| 2051 | Blood Pressure - Both | 723237002 | Non-invasive blood pressure (observable entity) |
| 2051 | Blood Pressure - Both | 1036531000000108 | Non-invasive central blood pressure (observable entity) |
| 2051 | Blood Pressure - Both | 170605004 | Normal blood pressure (finding) |
| 2051 | Blood Pressure - Both | 2004005 | Normal blood pressure (finding) |
| 2051 | Blood Pressure - Both | 147851006 | Normal blood pressure (finding) |
| 2051 | Blood Pressure - Both | 140249002 | O/E - blood pressure decreased (finding) |
| 2051 | Blood Pressure - Both | 140233004 | O/E - blood pressure reading (finding) |
| 2051 | Blood Pressure - Both | 140237003 | O/E - BP borderline low (finding) |
| 2051 | Blood Pressure - Both | 140239000 | O/E - BP borderline raised (finding) |
| 2051 | Blood Pressure - Both | 140236007 | O/E - BP reading low (finding) |
| 2051 | Blood Pressure - Both | 140238008 | O/E - BP reading normal (finding) |
| 2051 | Blood Pressure - Both | 140240003 | O/E - BP reading raised (finding) |
| 2051 | Blood Pressure - Both | 140241004 | O/E - BP reading very high (finding) |
| 2051 | Blood Pressure - Both | 140235006 | O/E - BP reading very low (finding) |
| 2051 | Blood Pressure - Both | 140253000 | O/E - BP reading: no postural drop (finding) |
| 2051 | Blood Pressure - Both | 163040000 | O/E - BP reading: no postural drop (finding) |
| 2051 | Blood Pressure - Both | 140242006 | O/E - BP reading:postural drop (finding) |
| 2051 | Blood Pressure - Both | 140245008 | O/E - BP stable (finding) |

| | | | |
|---|---|---|---|
| 2051 | Blood Pressure - Both | 147835003 | O/E - initial high BP (finding) |
| 2051 | Blood Pressure - Both | 140256008 | O/E-blood pressure reading NOS (finding) |
| 2051 | Blood Pressure - Both | 163024003 | On examination - blood pressure borderline low (disorder) |
| 2051 | Blood Pressure - Both | 163026001 | On examination - blood pressure borderline raised (finding) |
| 2051 | Blood Pressure - Both | 163036009 | On examination - blood pressure decreased (finding) |
| 2051 | Blood Pressure - Both | 163020007 | On examination - blood pressure reading (finding) |
| 2051 | Blood Pressure - Both | 163023009 | On examination - blood pressure reading low (disorder) |
| 2051 | Blood Pressure - Both | 163025002 | On examination - blood pressure reading normal (finding) |
| 2051 | Blood Pressure - Both | 163043003 | On examination - blood pressure reading NOS (finding) |
| 2051 | Blood Pressure - Both | 532221000000107 | On examination - blood pressure reading NOS (finding) |
| 2051 | Blood Pressure - Both | 163027005 | On examination - blood pressure reading raised (finding) |
| 2051 | Blood Pressure - Both | 163028000 | On examination - blood pressure reading very high (finding) |
| 2051 | Blood Pressure - Both | 163022004 | On examination - blood pressure reading very low (disorder) |
| 2051 | Blood Pressure - Both | 313005002 | On examination - blood pressure reading: no postural drop (situation) |
| 2051 | Blood Pressure - Both | 163029008 | On examination - blood pressure reading: postural drop (finding) |
| 2051 | Blood Pressure - Both | 163032006 | On examination - blood pressure stable (finding) |
| 2051 | Blood Pressure - Both | 170581003 | On examination - initial high blood pressure (finding) |
| 2051 | Blood Pressure - Both | 271647008 | Raised blood pressure (finding) |
| 2051 | Blood Pressure - Both | 271869003 | Raised blood pressure reading (disorder) |
| 2051 | Blood Pressure - Both | 337061000000109 | Self measured blood pressure reading |
| 2051 | Blood Pressure - Both | 337501000000104 | Self measured blood pressure reading |
| 2051 | Blood Pressure - Both | 335661000000109 | Self measured blood pressure reading (observable entity) |
| 2051 | Blood Pressure - Both | 163034007 | Standing blood pressure (observable entity) |
| 2051 | Blood Pressure - Both | 140247000 | Standing blood pressure reading (observable entity) |
| 2051 | Blood Pressure - Both | 364090009 | Systemic arterial pressure (observable entity) |
| 2051 | Blood Pressure - Both | 386536003 | Systemic blood pressure (observable entity) |
| 2052 | Blood Pressure - Diastolic | 213051000000103 | Ambulatory diastolic blood pressure |
| 2052 | Blood Pressure - Diastolic | 213061000000100 | Ambulatory diastolic blood pressure |
| 2052 | Blood Pressure - Diastolic | 198091000000104 | Ambulatory diastolic blood pressure (observable entity) |
| 2052 | Blood Pressure - Diastolic | 167061000000106 | Average 24 hour diastolic blood pressure |
| 2052 | Blood Pressure - Diastolic | 165011000000100 | Average 24 hour diastolic blood pressure |
| 2052 | Blood Pressure - Diastolic | 314462001 | Average 24 hour diastolic blood pressure (observable entity) |
| 2052 | Blood Pressure - Diastolic | 165031000000108 | Average day interval diastolic blood pressure |
| 2052 | Blood Pressure - Diastolic | 167081000000102 | Average day interval diastolic blood pressure |
| 2052 | Blood Pressure - Diastolic | 314461008 | Average day interval diastolic blood pressure (observable entity) |
| 2052 | Blood Pressure - Diastolic | 918051000000106 | Average diastolic blood pressure |
| 2052 | Blood Pressure - Diastolic | 314453003 | Average diastolic blood pressure (observable entity) |
| 2052 | Blood Pressure - Diastolic | 167121000000104 | Average home diastolic blood pressure |
| 2052 | Blood Pressure - Diastolic | 165071000000105 | Average home diastolic blood pressure |
| 2052 | Blood Pressure - Diastolic | 413605002 | Average home diastolic blood pressure (observable entity) |
| 2052 | Blood Pressure - Diastolic | 165051000000101 | Average night interval diastolic blood pressure |
| 2052 | Blood Pressure - Diastolic | 167101000000108 | Average night interval diastolic blood pressure |
| 2052 | Blood Pressure - Diastolic | 314460009 | Average night interval diastolic blood pressure (observable entity) |
| 2052 | Blood Pressure - Diastolic | 945861000000100 | Baseline diastolic blood pressure |
| 2052 | Blood Pressure - Diastolic | 945851000000103 | Baseline diastolic blood pressure (observable entity) |
| 2052 | Blood Pressure - Diastolic | 716632005 | Baseline diastolic blood pressure (observable entity) |

| | | | |
|---|---|---|---|
| 2052 | Blood Pressure - Diastolic | 1036581000000107 | Central diastolic blood pressure |
| 2052 | Blood Pressure - Diastolic | 1091811000000102 | Diastolic arterial pressure (observable entity) |
| 2052 | Blood Pressure - Diastolic | 67726005 | Diastolic arterial pressure (observable entity) |
| 2052 | Blood Pressure - Diastolic | 271650006 | Diastolic blood pressure (observable entity) |
| 2052 | Blood Pressure - Diastolic | 446226005 | Diastolic blood pressure on admission (observable entity) |
| 2052 | Blood Pressure - Diastolic | 251073000 | Invasive diastolic arterial pressure (observable entity) |
| 2052 | Blood Pressure - Diastolic | 95311000000107 | Lying diastolic blood pressure |
| 2052 | Blood Pressure - Diastolic | 100181000000103 | Lying diastolic blood pressure |
| 2052 | Blood Pressure - Diastolic | 407557002 | Lying diastolic blood pressure (observable entity) |
| 2052 | Blood Pressure - Diastolic | 1036571000000105 | Non-invasive central diastolic blood pressure (observable entity) |
| 2052 | Blood Pressure - Diastolic | 174255007 | Non-invasive diastolic arterial pressure (observable entity) |
| 2052 | Blood Pressure - Diastolic | 251072005 | Non-invasive diastolic arterial pressure (observable entity) |
| 2052 | Blood Pressure - Diastolic | 140244007 | O/E - Diastolic BP reading (finding) |
| 2052 | Blood Pressure - Diastolic | 163031004 | On examination - Diastolic blood pressure reading (finding) |
| 2052 | Blood Pressure - Diastolic | 95291000000106 | Sitting diastolic blood pressure |
| 2052 | Blood Pressure - Diastolic | 100161000000107 | Sitting diastolic blood pressure |
| 2052 | Blood Pressure - Diastolic | 407555005 | Sitting diastolic blood pressure (observable entity) |
| 2052 | Blood Pressure - Diastolic | 75141000000102 | Standing diastolic blood pressure |
| 2052 | Blood Pressure - Diastolic | 79411000000108 | Standing diastolic blood pressure |
| 2052 | Blood Pressure - Diastolic | 400975005 | Standing diastolic blood pressure (observable entity) |
| 2053 | Blood Pressure - Systolic | 314464000 | 24 hour systolic blood pressure (observable entity) |
| 2053 | Blood Pressure - Systolic | 213031000000105 | Ambulatory systolic blood pressure |
| 2053 | Blood Pressure - Systolic | 213041000000101 | Ambulatory systolic blood pressure |
| 2053 | Blood Pressure - Systolic | 198081000000101 | Ambulatory systolic blood pressure (observable entity) |
| 2053 | Blood Pressure - Systolic | 167071000000104 | Average 24 hour systolic blood pressure |
| 2053 | Blood Pressure - Systolic | 165021000000106 | Average 24 hour systolic blood pressure |
| 2053 | Blood Pressure - Systolic | 314449000 | Average 24 hour systolic blood pressure (observable entity) |
| 2053 | Blood Pressure - Systolic | 165041000000104 | Average day interval systolic blood pressure |
| 2053 | Blood Pressure - Systolic | 167091000000100 | Average day interval systolic blood pressure |
| 2053 | Blood Pressure - Systolic | 314446007 | Average day interval systolic blood pressure (observable entity) |
| 2053 | Blood Pressure - Systolic | 165081000000107 | Average home systolic blood pressure |
| 2053 | Blood Pressure - Systolic | 167131000000102 | Average home systolic blood pressure |
| 2053 | Blood Pressure - Systolic | 413606001 | Average home systolic blood pressure (observable entity) |
| 2053 | Blood Pressure - Systolic | 165061000000103 | Average night interval systolic blood pressure |
| 2053 | Blood Pressure - Systolic | 167111000000105 | Average night interval systolic blood pressure |
| 2053 | Blood Pressure - Systolic | 314445006 | Average night interval systolic blood pressure (observable entity) |
| 2053 | Blood Pressure - Systolic | 918041000000108 | Average systolic blood pressure |
| 2053 | Blood Pressure - Systolic | 314440001 | Average systolic blood pressure (observable entity) |
| 2053 | Blood Pressure - Systolic | 945881000000109 | Baseline systolic blood pressure |
| 2053 | Blood Pressure - Systolic | 945871000000107 | Baseline systolic blood pressure (observable entity) |
| 2053 | Blood Pressure - Systolic | 716579001 | Baseline systolic blood pressure (observable entity) |
| 2053 | Blood Pressure - Systolic | 1036561000000103 | Central systolic blood pressure |
| 2053 | Blood Pressure - Systolic | 251071003 | Invasive systolic arterial pressure (observable entity) |
| 2053 | Blood Pressure - Systolic | 100171000000100 | Lying systolic blood pressure |
| 2053 | Blood Pressure - Systolic | 95301000000105 | Lying systolic blood pressure |
| 2053 | Blood Pressure - Systolic | 407556006 | Lying systolic blood pressure (observable entity) |

| | | | |
|---|---|---|---|
| 2053 | Blood Pressure - Systolic | 314448008 | Maximum 24 hour systolic blood pressure (observable entity) |
| 2053 | Blood Pressure - Systolic | 314444005 | Maximum day interval systolic blood pressure (observable entity) |
| 2053 | Blood Pressure - Systolic | 314443004 | Maximum night interval systolic blood pressure (observable entity) |
| 2053 | Blood Pressure - Systolic | 314439003 | Maximum systolic blood pressure (observable entity) |
| 2053 | Blood Pressure - Systolic | 314447003 | Minimum 24 hour systolic blood pressure (observable entity) |
| 2053 | Blood Pressure - Systolic | 314441002 | Minimum day interval systolic blood pressure (observable entity) |
| 2053 | Blood Pressure - Systolic | 314442009 | Minimum night interval systolic blood pressure (observable entity) |
| 2053 | Blood Pressure - Systolic | 314438006 | Minimum systolic blood pressure (observable entity) |
| 2053 | Blood Pressure - Systolic | 1036551000000101 | Non-invasive central systolic blood pressure (observable entity) |
| 2053 | Blood Pressure - Systolic | 251070002 | Non-invasive systolic arterial pressure (observable entity) |
| 2053 | Blood Pressure - Systolic | 140243001 | O/E - Systolic BP reading (finding) |
| 2053 | Blood Pressure - Systolic | 163030003 | On examination - Systolic blood pressure reading (finding) |
| 2053 | Blood Pressure - Systolic | 1104341000000101 | Royal College of Physicians National Early Warning Score 2 - systolic blood pressure score (observable entity) |
| 2053 | Blood Pressure - Systolic | 95281000000109 | Sitting systolic blood pressure |
| 2053 | Blood Pressure - Systolic | 100151000000109 | Sitting systolic blood pressure |
| 2053 | Blood Pressure - Systolic | 407554009 | Sitting systolic blood pressure (observable entity) |
| 2053 | Blood Pressure - Systolic | 75131000000106 | Standing systolic blood pressure |
| 2053 | Blood Pressure - Systolic | 79401000000106 | Standing systolic blood pressure |
| 2053 | Blood Pressure - Systolic | 400974009 | Standing systolic blood pressure (observable entity) |
| 2053 | Blood Pressure - Systolic | 72313002 | Systolic arterial pressure (observable entity) |
| 2053 | Blood Pressure - Systolic | 271649006 | Systolic blood pressure (observable entity) |
| 2053 | Blood Pressure - Systolic | 708502007 | Systolic blood pressure of neonate at birth (observable entity) |
| 2053 | Blood Pressure - Systolic | 399304008 | Systolic blood pressure on admission (observable entity) |
| 2050 | HbA1c | 567641000000108 | Haemoglobin A1C - diabetic control NOS (observable entity) |
| 2050 | HbA1c | 936121000000108 | Haemoglobin A1c (diagnostic reference range) |
| 2050 | HbA1c | 1049311000000103 | Haemoglobin A1c (diagnostic reference range) - International Federation of Clinical Chemistry and Laboratory Medicine standardised |
| 2050 | HbA1c | 955411000000102 | oglobin A1c (diagnostic reference range) - International Federation of Clinical Chemistry and Laboratory Medicine standardised (observable entity) |
| 2050 | HbA1c | 936111000000102 | Haemoglobin A1c (diagnostic reference range) (observable entity) |
| 2050 | HbA1c | 1010951000000100 | Haemoglobin A1c (diagnostic reference range) (observable entity) |
| 2050 | HbA1c | 1049331000000106 | Haemoglobin A1c (monitoring ranges) - International Federation of Clinical Chemistry and Laboratory Medicine standardised |
| 2050 | HbA1c | 955421000000108 | aemoglobin A1c (monitoring ranges) - International Federation of Clinical Chemistry and Laboratory Medicine standardised (observable entity) |
| 2050 | HbA1c | 936131000000105 | Haemoglobin A1c (monitoring ranges) (observable entity) |
| 2050 | HbA1c | 1010941000000103 | Haemoglobin A1c (monitoring ranges) (observable entity) |
| 2050 | HbA1c | 936141000000101 | Haemoglobin A1c (monitoring reference ranges) |
| 2050 | HbA1c | 999791000000106 | Haemoglobin A1c level - International Federation of Clinical Chemistry and Laboratory Medicine standardised (observable entity) |
| 2050 | HbA1c | 1019431000000105 | Haemoglobin A1c level (Diabetes Control and Complications Trial aligned) (observable entity) |
| 2050 | HbA1c | 1049301000000100 | lobin A1c level (diagnostic reference range) - International Federation of Clinical Chemistry and Laboratory Medicine standardised (observable entity) |
| 2050 | HbA1c | 1049321000000109 | moglobin A1c level (monitoring ranges) - International Federation of Clinical Chemistry and Laboratory Medicine standardised (observable entity) |
| 2050 | HbA1c | 1003671000000109 | Haemoglobin A1c level (observable entity) |
| 2050 | HbA1c | 491841000000105 | Haemoglobin A1c level (observable entity) |
| 2050 | HbA1c | 727941000000108 | Haemoglobin A1c target - International Federation of Clinical Chemistry and Laboratory Medicine standardised |
| 2050 | HbA1c | 727931000000104 | Haemoglobin A1c target - International Federation of Clinical Chemistry and Laboratory Medicine standardised (observable entity) |
| 2050 | HbA1c | 165683005 | Hb. A1C - diabetic control NOS (observable entity) |
| 2050 | HbA1c | 143085005 | Hb. A1C - diabetic control NOS (procedure) |
| 2050 | HbA1c | 143080000 | Hb. A1C - glycated haemoglobin (& [diabetic control]) (procedure) |
| 2050 | HbA1c | 165678002 | Hb. A1C - glycated haemoglobin (& [diabetic control]) (procedure) |

| | | | |
|---|---|---|---|
| 2050 | HbA1c | 143127002 | HbA1 - diabetic control (observable entity) |
| 2050 | HbA1c | 165723000 | HbA1 - diabetic control (observable entity) |
| 2050 | HbA1c | 310309003 | HbA1 - diabetic control interpretation (observable entity) |
| 2050 | HbA1c | 86551000000108 | HBA1c target |
| 2050 | HbA1c | 107991000000100 | HBA1c target |
| 2050 | HbA1c | 269823000 | Hemoglobin A1C - diabetic control interpretation (observable entity) |
| 2050 | HbA1c | 408591000 | Hemoglobin A1c target (observable entity) |
| 2050 | HbA1c | 446074002 | noglobin A1c target value using International Federation of Clinical Chemistry and Laboratory Medicine standardized method (observable entity) |
| 2050 | HbA1c | 443911005 | Ordinal level of hemoglobin A1c (observable entity) |
| 2050 | HbA1c | 1107481000000106 | Substance concentration of haemoglobin A1c in blood (observable entity) |
| 2050 | HbA1c | 1013511000000100 | Total glycosylated haemoglobin level (observable entity) |
| 1523 | HbA1c_DCCT | 1019431000000105 | Haemoglobin A1c level (Diabetes Control and Complications Trial aligned) (observable entity) |
| 1522 | HbA1c_IFCC | 936121000000108 | Haemoglobin A1c (diagnostic reference range) |
| 1522 | HbA1c_IFCC | 1049311000000103 | Haemoglobin A1c (diagnostic reference range) - International Federation of Clinical Chemistry and Laboratory Medicine standardised |
| 1522 | HbA1c_IFCC | 955411000000102 | oglobin A1c (diagnostic reference range) - International Federation of Clinical Chemistry and Laboratory Medicine standardised (observable entity) |
| 1522 | HbA1c_IFCC | 936111000000102 | Haemoglobin A1c (diagnostic reference range) (observable entity) |
| 1522 | HbA1c_IFCC | 1049331000000106 | Haemoglobin A1c (monitoring ranges) - International Federation of Clinical Chemistry and Laboratory Medicine standardised |
| 1522 | HbA1c_IFCC | 955421000000108 | aemoglobin A1c (monitoring ranges) - International Federation of Clinical Chemistry and Laboratory Medicine standardised (observable entity) |
| 1522 | HbA1c_IFCC | 936131000000105 | Haemoglobin A1c (monitoring ranges) (observable entity) |
| 1522 | HbA1c_IFCC | 936141000000101 | Haemoglobin A1c (monitoring reference ranges) |
| 1522 | HbA1c_IFCC | 999791000000106 | Haemoglobin A1c level - International Federation of Clinical Chemistry and Laboratory Medicine standardised (observable entity) |
| 1522 | HbA1c_IFCC | 1049301000000100 | lobin A1c level (diagnostic reference range) - International Federation of Clinical Chemistry and Laboratory Medicine standardised (observable entity) |
| 1522 | HbA1c_IFCC | 1049321000000109 | noglobin A1c level (monitoring ranges) - International Federation of Clinical Chemistry and Laboratory Medicine standardised (observable entity) |
| 1524 | HbA1c_Unspecified | 1010951000000100 | Haemoglobin A1c (diagnostic reference range) (observable entity) |
| 1524 | HbA1c_Unspecified | 1010941000000103 | Haemoglobin A1c (monitoring ranges) (observable entity) |
| 1524 | HbA1c_Unspecified | 1003671000000109 | Haemoglobin A1c level (observable entity) |
| 1524 | HbA1c_Unspecified | 1107481000000106 | Substance concentration of haemoglobin A1c in blood (observable entity) |
| 1518 | INRMonitor_num | 142988001 | International normalised ratio (procedure) |
| 1518 | INRMonitor_num | 165581004 | International normalized ratio (observable entity) |
| 1518 | INRMonitor_num | 440432009 | International normalized ratio result obtained using portable international normalized ratio monitoring device (observable entity) |
| 2056 | Total cholesterol | 84698008 | Cholesterol (substance) |
| 2056 | Total cholesterol | 18520611000001106 | Cholesterol (substance) |
| 2056 | Total cholesterol | 301875003 | Cholesterol analyte (substance) |
| 2056 | Total cholesterol | 850981000000101 | Cholesterol level (observable entity) |
| 2056 | Total cholesterol | 852401000000104 | Maximum cholesterol level (observable entity) |
| 2056 | Total cholesterol | 852411000000102 | Minimum cholesterol level (observable entity) |
| 2056 | Total cholesterol | 1017161000000104 | Plasma total cholesterol level (observable entity) |
| 2056 | Total cholesterol | 1005671000000105 | Serum cholesterol level (observable entity) |
| 2056 | Total cholesterol | 1026441000000100 | Serum cholesterol studies (observable entity) |
| 2056 | Total cholesterol | 1083761000000106 | Serum fasting total cholesterol level (observable entity) |
| 2056 | Total cholesterol | 994351000000103 | Serum total cholesterol level (observable entity) |
| 2056 | Total cholesterol | 1106531000000105 | Substance concentration of cholesterol in plasma (observable entity) |
| 2056 | Total cholesterol | 1106541000000101 | Substance concentration of cholesterol in serum (observable entity) |
| 2056 | Total cholesterol | 301860006 | Total cholesterol (substance) |
| 2056 | Total cholesterol | 853681000000104 | Total cholesterol level (observable entity) |
